# An observational cohort study on the incidence of SARS-CoV-2 infection and B.1.1.7 variant infection in healthcare workers by antibody and vaccination status

**DOI:** 10.1101/2021.03.09.21253218

**Authors:** Sheila F Lumley, Gillian Rodger, Bede Constantinides, Nicholas Sanderson, Kevin K Chau, Teresa L Street, Denise O’Donnell, Alison Howarth, Stephanie B Hatch, Brian D Marsden, Stuart Cox, Tim James, Fiona Warren, Liam J Peck, Thomas G Ritter, Zoe de Toledo, Laura Warren, David Axten, Richard J Cornall, E Yvonne Jones, David I Stuart, Gavin Screaton, Daniel Ebner, Sarah Hoosdally, Meera Chand, Oxford University Hospitals Staff Testing Group, Derrick W Crook, Anne-Marie O’Donnell, Christopher P Conlon, Koen B Pouwels, A Sarah Walker, Tim EA Peto, Susan Hopkins, Timothy M Walker, Nicole E Stoesser, Philippa C Matthews, Katie Jeffery, David W Eyre

**Affiliations:** Oxford University Hospitals NHS Foundation Trust, Oxford, UK; Nuffield Department of Medicine, University of Oxford, Oxford, UK; NIHR Oxford Biomedical Research Centre, University of Oxford, Oxford, UK; NIHR Health Protection Research Unit in Healthcare Associated Infections and Antimicrobial Resistance at University of Oxford in partnership with Public Health England, Oxford, UK; Kennedy Institute of Rheumatology Research, University of Oxford, UK; Medical School, University of Oxford, Oxford, UK; Target Discovery Institute, University of Oxford, Oxford, UK; National Infection Service, Public Health England Colindale, UK; Nuffield Department of Population Health, University of Oxford, Oxford, UK; Oxford University Clinical Research Unit, Ho Chi Minh City, Vietnam; Big Data Institute, University of Oxford, Oxford, UK

**Keywords:** SARS-CoV-2, vaccine, antibody, healthcare worker, immunity

## Abstract

**Background:** Natural and vaccine-induced immunity will play a key role in controlling the SARS-CoV-2 pandemic. SARS-CoV-2 variants have the potential to evade natural and vaccine-induced immunity.

**Methods:** In a longitudinal cohort study of healthcare workers (HCWs) in Oxfordshire, UK, we investigated the protection from symptomatic and asymptomatic PCR-confirmed SARS-CoV-2 infection conferred by vaccination (Pfizer-BioNTech BNT162b2, Oxford-AstraZeneca ChAdOx1 nCOV-19) and prior infection (determined using anti-spike antibody status), using Poisson regression adjusted for age, sex, temporal changes in incidence and role. We estimated protection conferred after one versus two vaccinations and from infections with the B.1.1.7 variant identified using whole genome sequencing.

**Results:** 13,109 HCWs participated; 8285 received the Pfizer-BioNTech vaccine (1407 two doses) and 2738 the Oxford-AstraZeneca vaccine (49 two doses). Compared to unvaccinated seronegative HCWs, natural immunity and two vaccination doses provided similar protection against symptomatic infection: no HCW vaccinated twice had symptomatic infection, and incidence was 98% lower in seropositive HCWs (adjusted incidence rate ratio 0.02 [95%CI <0.01-0.18]). Two vaccine doses or seropositivity reduced the incidence of any PCR-positive result with or without symptoms by 90% (0.10 [0.02-0.38]) and 85% (0.15 [0.08-0.26]) respectively. Single-dose vaccination reduced the incidence of symptomatic infection by 67% (0.33 [0.21-0.52]) and any PCR-positive result by 64% (0.36 [0.26-0.50]). There was no evidence of differences in immunity induced by natural infection and vaccination for infections with S-gene target failure and B.1.1.7.

**Conclusion:** Natural infection resulting in detectable anti-spike antibodies and two vaccine doses both provide robust protection against SARS-CoV-2 infection, including against the B.1.1.7 variant.

**Summary:** Natural infection resulting in detectable anti-spike antibodies and two vaccine doses both provided ≥ 85% protection against symptomatic and asymptomatic SARS-CoV-2 infection in healthcare workers, including against the B.1.1.7 variant. Single dose vaccination reduced symptomatic infection by 67%.

## Introduction

The SARS-CoV-2 pandemic has had a global impact on morbidity and mortality.^1^ Natural and vaccine-induced immunity will play a key role in controlling the pandemic, by reducing transmission, hospitalisation and mortality. However, the ability of new SARS-CoV-2 variants to evade natural and vaccine-induced immunity mounted against ancestral viruses is of major public health concern.

Prior SARS-CoV-2 infection protects against PCR-confirmed symptomatic/asymptomatic SARS-CoV-2 infection by 83-88% up to 5-6 months, with greater reductions in symptomatic infections.^2–4^ Ongoing longitudinal studies are required to determine the duration of protection conferred by natural immunity; however evaluating this will be more difficult with widespread vaccination. Understanding the interaction between prior infection/serostatus and vaccination on protection from infection is also important.

Three vaccines have been approved for use in the UK to date^5^, with Pfizer-BioNTech BNT162b2 and Oxford-AstraZeneca ChAdOx1 nCoV-19 (AZD1222) currently the most widely deployed, with many individuals receiving only one dose to date following a Government decision to extend the dosing interval to 12 weeks to maximise initial coverage. For BNT162b2, trials demonstrated 95% efficacy in preventing symptomatic PCR-confirmed infection >7 days post-second dose; these findings have been replicated in several real-world studies including in Israel (92% effectiveness)^6^ and the UK (88% effectiveness in individuals >80 years^7^; 85% reduction in all-PCR positives in a cohort of healthcare workers [HCWs]).^8^ Vaccine efficacy of 50-90% is seen following a single dose, dependent on population demographics, exposures and time-frame studied.^6,7,9–13^ Fewer real-world data are available for ChAdOx1 nCoV-19, due to its later regulatory approval. Trials demonstrated vaccine efficacy of 62% against PCR-positive infection >14 days post-second dose using a standard dose/standard dose regimen, with subsequent analysis showing a higher efficacy of 81% in those with a longer dosing interval (>12 weeks). Single dose vaccine efficacy >22 days post-first dose has been reported as 69-76%.^14,15^ No real-world data on vaccine effectiveness against PCR-positive infections has been published, but preliminary analyses show a reduction in hospital admissions in Scotland.^16^

A novel SARS-CoV-2 variant, B.1.1.7, identified in September-2020 in the UK, has spread rapidly. Estimates suggest increased transmissibility and disease severity.^17–20^ The lineage carries several mutations of immunologic significance, including N501Y located in the receptor-binding domain (RBD), a key neutralising antibody target; deletions in the N-terminal domain at residues 69/70, associated with viral escape in the immunocompromised and S-gene target failure (SGTF) in PCR assays; and a deletion at residue 144 resulting in decreased monoclonal antibody binding.^21^

Reinfection rates following natural infection have not been shown to be higher in studies using SGTF as a proxy for B.1.1.7,^20,22^ even though variably decreased sensitivity to neutralisation by monoclonal antibodies, convalescent plasma and sera from vaccinated individuals has been observed *in vitro* for B.1.1.7.^23–34^ The Oxford-AstraZeneca trial showed good vaccine efficacy against sequencing-confirmed symptomatic B.1.1.7, despite evidence of decreased neutralising titres, but decreased efficacy for asymptomatic/unknown symptom infections.^35^ Pfizer-BioNTech vaccine effectiveness in HCWs appears preserved despite increasing B.1.1.7 incidence in the UK; however these studies have not specifically investigated cases of SGTF or sequencing-confirmed B.1.1.7.^8,36^

We use an observational longitudinal cohort study of SARS-CoV-2 infection in hospital HCWs to investigate and compare the protection from SARS-CoV-2 infection conferred by vaccination and prior infection (determined using anti-spike antibody status). Additionally, we estimate the protection conferred by different vaccines, after one versus two doses and from infections with the B.1.1.7 variant confirmed by whole-genome sequencing.

## Methods

### Setting

Oxford University Hospitals (OUH) offers symptomatic and asymptomatic SARS-CoV-2 testing to all staff at four hospitals and associated facilities in Oxfordshire, UK. SARS-CoV-2 PCR testing of nasal and oropharyngeal swabs for symptomatic (new persistent cough, fever ≥ 37.8°C, anosmia/ageusia) staff was offered from 27-March-2020. Asymptomatic HCWs were offered voluntary nasal and oropharyngeal swab PCR testing every two weeks and serological testing every two months from 23-April-2020, as previously described.^2,37,38^ We report data up to 28-February-2021. To minimise under-ascertainment of outcomes arising from staff leaving OUH’s employment, only those who participated in asymptomatic screening, symptomatic testing or vaccination from 01-September-2020 onwards were included. We also performed a sensitivity analysis restricted to staff participating in asymptomatic screening or symptomatic testing from 01-September-2020. There was no limit on study size; all staff working for the hospitals were eligible to participate.

### Laboratory assays

Antibody status was determined using an anti-trimeric spike IgG ELISA^39^ using an 8 million units threshold to determine antibody-positivity. PCR tests were performed by OUH using a range of PCR assays (see Supplement). PCR-positive results from symptomatic community testing were also recorded. From 16-November-2020, OUH used the Thermo Fisher TaqPath PCR assay as their first-line diagnostic assay, which includes orf1ab, S and N gene targets. As such SGTF indicative of the B.1.1.7 variant^20^ could be identified, i.e. orf1ab-positive/N-positive only. Oxford Nanopore sequencing was undertaken of all stored PCR-positive primary samples from 01-December-2020 onwards to identify the infecting lineage (see Supplement).

### Study groups

Staff members were classified into five groups: a) unvaccinated and consistently seronegative during follow-up; b) unvaccinated and ever seropositive; c) vaccinated once, always seronegative prior to vaccination; d) vaccinated twice, always seronegative prior to first vaccination; e) vaccinated (once or twice) and ever seropositive prior to first vaccination. The latter group were combined as relatively few staff were previously seropositive and vaccinated twice. Vaccinated groups were considered at-risk of infection >14 days after each vaccine dose (see Table 1 for further details of at-risk periods).

**Table 1.**
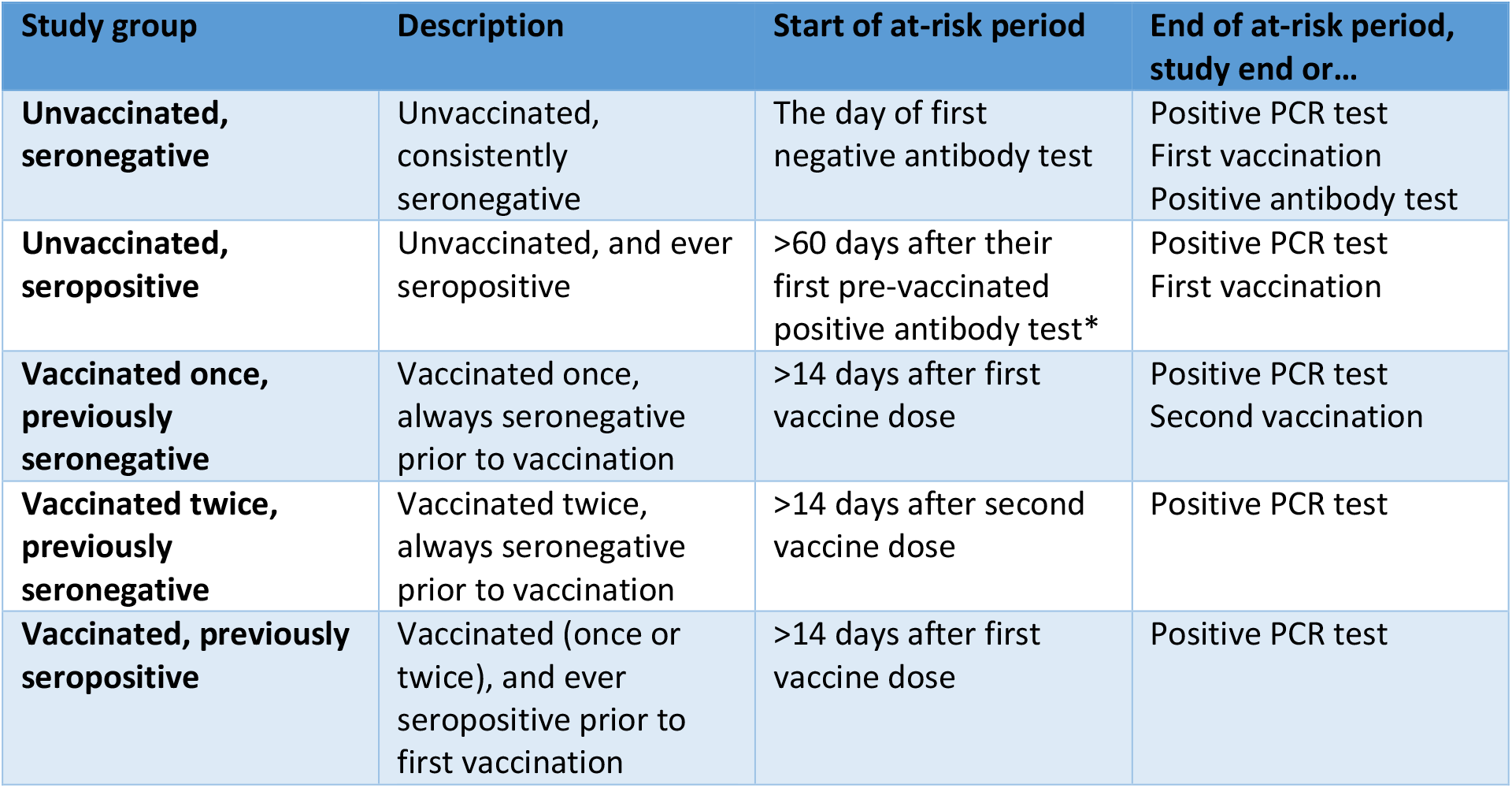
Study follow up groups. *To allow for any persistent RNA from the first infection and also requiring >60 days since the last positive PCR test. Those who were vaccinated without any prior antibody measurement were included in the previously seronegative follow up groups.

| Study group | Description | Start of at-risk period | End of at-risk period, study end or... |
| --- | --- | --- | --- |
| **Unvaccinated, seronegative** | Unvaccinated, consistently seronegative | The day of first negative antibody test | Positive PCR test<br>First vaccination<br>Positive antibody test |
| **Unvaccinated, seropositive** | Unvaccinated, and ever seropositive | >60 days after their first pre-vaccinated positive antibody test* | Positive PCR test<br>First vaccination |
| **Vaccinated once, previously seronegative** | Vaccinated once, always seronegative prior to vaccination | >14 days after first vaccine dose | Positive PCR test<br>Second vaccination |
| **Vaccinated twice, previously seronegative** | Vaccinated twice, always seronegative prior to vaccination | >14 days after second vaccine dose | Positive PCR test |
| **Vaccinated, previously seropositive** | Vaccinated (once or twice), and ever seropositive prior to first vaccination | >14 days after first vaccine dose | Positive PCR test |

Staff remained at risk of infection in each follow-up group until the earliest of the study end, first vaccination, second vaccination in previously seronegative HCWs, a positive PCR test, or for unvaccinated HCWs, a positive antibody test. Staff could transition from one group to another following seroconversion or vaccination after 60 or 14 days respectively, disregarding any PCR-positive result during this cross-over period, including the 14 days following a second vaccination for previously seronegative HCWs vaccinated twice.

The staff vaccination programme began on 8-December-2020, starting with the Pfizer-BioNTech BNT162b2 vaccine, with the addition of the Oxford-AstraZeneca ChAdOx1 nCoV-19 vaccine from 4-January-2021. Some staff members received the ChAdOx1 nCoV-19 vaccine in clinical trials beginning 23-April-2020 and were included following unblinding.

### Outcomes

The main outcome was PCR-confirmed symptomatic SARS-CoV-2 infection. We also considered any PCR-positive result (i.e. either symptomatic or asymptomatic). To assess the impact of the B.1.1.7 variant on (re)infection risk, we also analysed PCR-positive results with and without SGTF, and those confirmed as B.1.1.7 on sequencing.

### Statistical analysis

We used Poisson regression to model incidence of each outcome per day-at-risk by study group. We adjusted for calendar month, age, sex, self-reported ethnicity and staff occupational role, patient contact and working on a non-ICU ward caring for Covid patients (previously shown to increase risk^37^) (details in Supplement). We compared incidence in each follow-up group to unvaccinated seronegative HCWs, using incidence rate ratios (IRRs), such that 100*(1-IRR) is the percentage protection arising from being seropositive or vaccinated. We tested for heterogeneity by vaccine type. To assess timing of onset of protection we also fitted models in vaccinated individuals from day 1 post-vaccination.

We used stacked Poisson regression to test for variation in the incidence of SGTF vs. non-SGTF PCR-positive results, and B.1.1.7 vs. non-B.1.1.7, considering only results from 01-December-2020 where S-gene PCR and sequencing were most complete.

We compared PCR cycle threshold (Ct) values between symptomatic and asymptomatic infections and by follow-up group using quantile regression.

## Ethics statement

Deidentified data were obtained from the Infections in Oxfordshire Research Database which has generic Research Ethics Committee, Health Research Authority and Confidentiality Advisory Group approvals (19/SC/0403, 19/CAG/0144).

## Results

13,109 individual HCWs contributed 2,835,260 person-days follow-up. 9765(74%) were female, the most common occupational roles were nurse (3579,27%), doctor (1776,14%), administrative staff (1688,13%) and healthcare assistant (1263,10%). The median(IQR) age was 39(30-50) years. Most HCWs were followed before vaccination: 10,513 HCWs were SARS-CoV-2 anti-spike IgG seronegative (2,274,675 person-days follow-up) and 1273 were seropositive (198,520 person-days). Most HCWs were vaccinated between December-2020 and January-2021 (Figure 1A); 8285 staff received Pfizer-BioNTech vaccine (1407 two doses) and 2738 Oxford-AstraZeneca vaccine (49 two doses). 11 HCWs received another vaccine or could not recall the manufacturer. Staff could move between follow-up groups; in total there were 9711 and 940 previously seronegative HCWs followed after a 1^st^ (289,134 person-days) and 2^nd^ (39,222 person-days) vaccine dose respectively, and 974 (33,709 person-days) in the vaccinated previously seropositive group.

**Figure 1.**
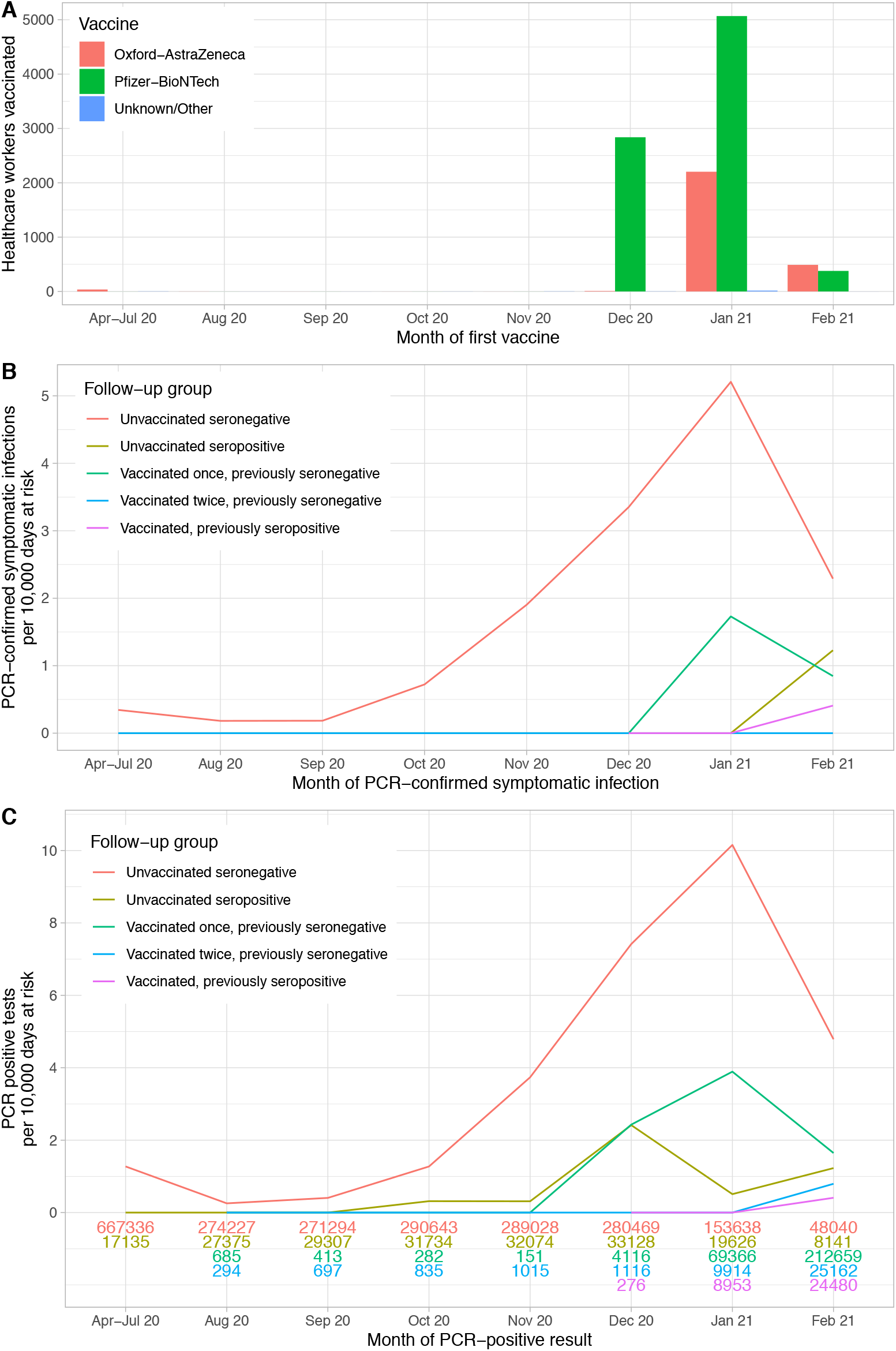
Vaccination timings (panel A) and observed incidence of symptomatic PCR-confirmed SARS-CoV-2 infection (panel B) and any PCR-positive result (panel C) by antibody and vaccine status. Some staff members received the Oxford-AstraZeneca vaccine in clinical trials beginning 23-April-2020 and were included following unblinding if in the active arm. The number of days at risk per month for each follow-up group is shown at the bottom of panel C. Due to small numbers rates are not plotted for vaccinated individuals prior to August 2020.

As previously reported,^2^ asymptomatic testing was less frequent in unvaccinated seropositive HCWs (127/10,000 person-days) than unvaccinated seronegative HCWs (185/10,000 person-days). Rates in previously seronegative and seropositive vaccinated staff were similar (163-169/10,000 person-days). Symptomatic testing followed a similar pattern (Table S1).

### Incidence of PCR-confirmed symptomatic SARS-CoV-2 infection

PCR-confirmed symptomatic SARS-CoV-2 infection in HCWs peaked in December-2020 and January-2021, similarly to local community-based infection rates^40^ (Figure 1B, Tables 2 and S2). 294 unvaccinated seronegative HCWs were infected, 1 unvaccinated seropositive HCW and 32 vaccinated HCWs >14 days post first vaccine (one previously seropositive). Compared to unvaccinated seronegative HCWs who had the highest rates of infection, incidence was 98% lower in unvaccinated seropositive HCWs (adjusted IRR [aIRR] 0.02 [95%CI <0.01-0.18; p<0.001]), and 67% lower following a first dose in previously seronegative HCWs (aIRR=0.33 [0.21-0.52; p<0.001]), with no symptomatic infections seen following a second dose (Figure 2). Incidence was also 93% lower in vaccinated previously seropositive HCWs (aIRR=0.07 [0.01-0.51; p=0.009]). Incidence was higher following a first vaccination than in seropositive HCWs (p=0.01), but there was no evidence of difference between seropositive HCWs and following a second vaccination (p=0.96). Independently of vaccination and antibody status, rates of infection were higher in staff caring for SARS-CoV-2-infected patients, in nurses and healthcare assistants, and in staff of Asian ethnicity (Table 2). Results from a sensitivity analysis restricting to only those participating in testing from 01-September-2020 were similar (n=11,758 HCWs, Table S3).

**Table 2.** Adjusted incidence rate ratios (IRRs) for symptomatic PCR-confirmed SARS-CoV-2 infection and any PCR-positive result (symptomatic or asymptomatic) by antibody and vaccine status. Event counts, follow-up, and unadjusted IRRs are provided in Table S2.

| Variable |  | Symptomatic PCR-confirmed infection |  |  | Any PCR-positive result |  |  |
| --- | --- | --- | --- | --- | --- | --- | --- |
|  |  | Adjusted IRR | 95% CI | p value | Adjusted IRR | 95% CI | p value |
| Age | Age, per 10 year increase | 0.92 | 0.84 - 1.02 | 0.10 | 0.99 | 0.99 - 1.00 | 0.07 |
| Sex | Female (reference) | 1.00 |  |  | 1.00 |  |  |
|  | Male | 1.11 | 0.85 - 1.44 | 0.46 | 1.08 | 0.90 - 1.31 | 0.41 |
| Patient facing role | No (reference) | 1.00 |  |  | 1.00 |  |  |
|  | Yes | 1.06 | 0.76 - 1.49 | 0.72 | 1.13 | 0.90 - 1.42 | 0.29 |
| Covid ward | Not working in Covid ward | 1.00 |  |  | 1.00 |  |  |
|  | Working in non-ICU Covid ward | 1.57 | 1.11 - 2.21 | 0.01 | 1.53 | 1.21 - 1.94 | <0.001 |
| Month | April - July 2020 (Reference) | 1.00 |  |  | 1.00 |  |  |
|  | August 2020 | 0.51 | 0.19 - 1.33 | 0.17 | 0.20 | 0.09 - 0.42 | <0.001 |
|  | September 2020 | 0.51 | 0.20 - 1.35 | 0.18 | 0.31 | 0.17 - 0.58 | <0.001 |
|  | October 2020 | 2.02 | 1.13 - 3.63 | 0.02 | 1.01 | 0.69 - 1.48 | 0.96 |
|  | November 2020 | 5.34 | 3.30 - 8.62 | <0.001 | 2.92 | 2.20 - 3.87 | <0.001 |
|  | December 2020 | 9.23 | 5.89 - 14.50 | <0.001 | 5.91 | 4.60 - 7.59 | <0.001 |
|  | January 2021 | 14.60 | 9.24 - 23.00 | <0.001 | 7.93 | 6.10 - 10.30 | <0.001 |
|  | February 2021 | 7.10 | 3.89 - 13.00 | <0.001 | 3.72 | 2.54 - 5.46 | <0.001 |
| Follow up group | Unvaccinated seronegative (Reference) | 1.00 |  |  | 1.00 |  |  |
|  | Unvaccinated seropositive | 0.02 | <0.01 - 0.18 | <0.001 | 0.15 | 0.08 - 0.26 | <0.001 |
|  | Vaccinated once, previously seronegative | 0.33 | 0.21 - 0.52 | <0.001 | 0.36 | 0.26 - 0.50 | <0.001 |
|  | Vaccinated twice, previously seronegative | No events |  |  | 0.10 | 0.02 - 0.38 | <0.001 |
|  | Vaccinated, previously seropositive | 0.07 | 0.01 - 0.51 | 0.009 | 0.04 | 0.01 - 0.27 | 0.001 |
| Ethnic group | White (Reference) | 1.00 |  |  | 1.00 |  |  |
|  | Asian | 1.90 | 1.47 - 2.46 | <0.001 | 1.59 | 1.32 - 1.91 | <0.001 |
|  | Black | 1.09 | 0.62 - 1.91 | 0.78 | 1.25 | 0.88 - 1.78 | 0.21 |
|  | Other | 1.31 | 0.87 - 1.97 | 0.20 | 1.23 | 0.93 - 1.62 | 0.16 |
| Role | Other (Reference) | 1.00 |  |  | 1.00 |  |  |
|  | Junior doctor | 1.35 | 0.86 - 2.12 | 0.20 | 1.10 | 0.78 - 1.54 | 0.59 |
|  | Senior doctor (Consultant) | 0.50 | 0.24 - 1.05 | 0.07 | 0.60 | 0.38 - 0.95 | 0.03 |
|  | Healthcare assistant | 1.71 | 1.18 - 2.47 | 0.005 | 1.82 | 1.42 - 2.34 | <0.001 |
|  | Nurse | 1.50 | 1.11 - 2.03 | 0.009 | 1.49 | 1.21 - 1.83 | <0.001 |
|  | Physio-, occupational or speech/language therapist | 0.65 | 0.26 - 1.61 | 0.35 | 1.17 | 0.73 - 1.88 | 0.51 |
|  | Porter, domestic staff | 1.05 | 0.48 - 2.29 | 0.91 | 1.37 | 0.84 - 2.22 | 0.20 |
|  | Administrator | 0.99 | 0.64 - 1.52 | 0.96 | 1.18 | 0.89 - 1.57 | 0.25 |

**Figure 2.**
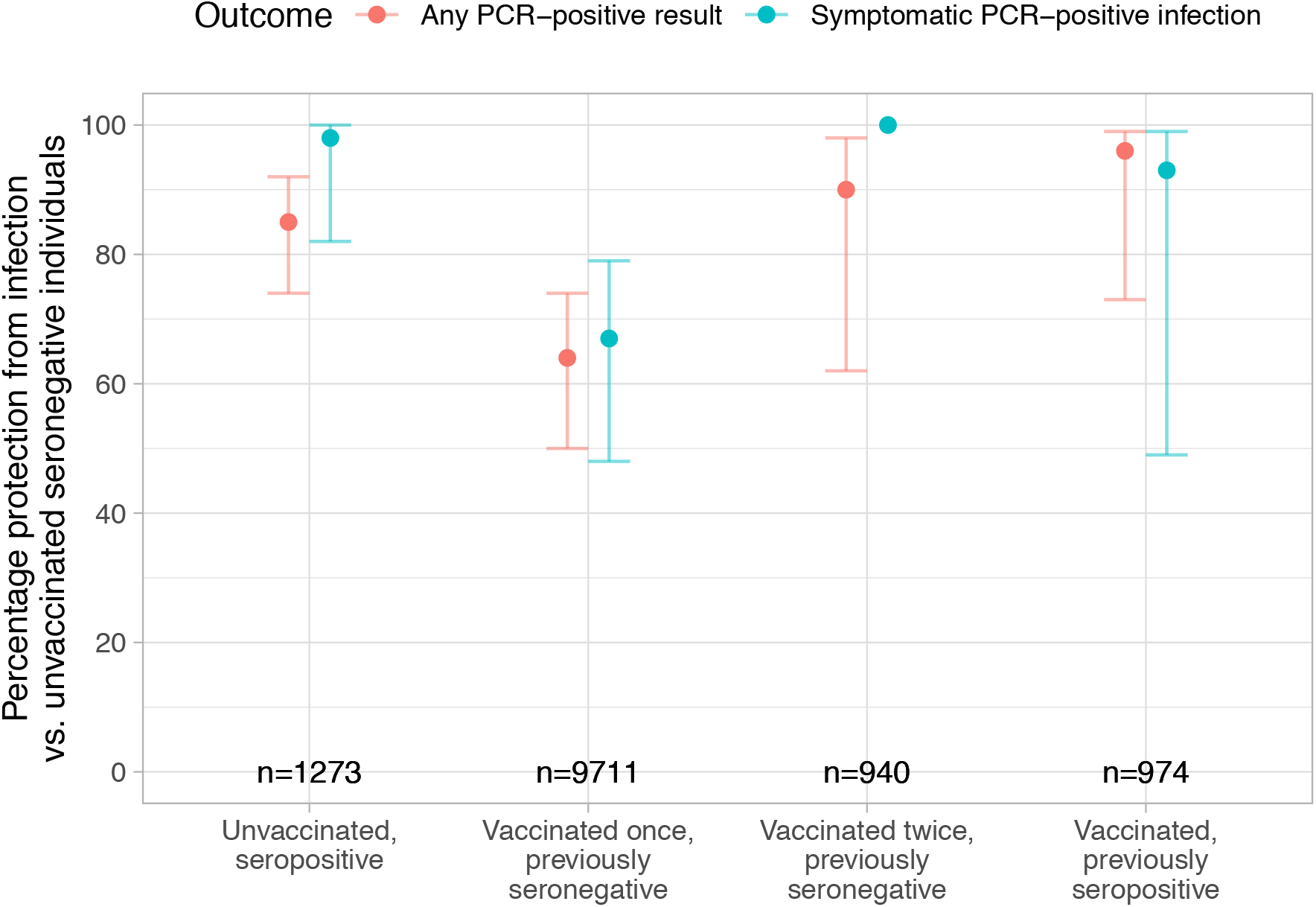
Protection from infection by antibody and vaccination status, compared with unvaccinated seronegative individuals. The number of HCWs in each follow up group is shown. 95% confidence intervals are plotted, except for previously seronegative HCWs vaccinated twice who had no symptomatic PCR confirmed infections.

38 unvaccinated seronegative HCWs attended hospital within −2 to +28 days of a SARS-CoV-2 PCR-positive result (14.2/million person-days); of these 27 had a Covid-19 primary diagnostic code and 16 required admission for Covid-19. Two previously seronegative vaccinated HCWs required hospital review (6.9/million person-days), however neither required admission. No HCW vaccinated twice or unvaccinated seropositive HCW required hospital review or admission.

### Incidence of any PCR-confirmed symptomatic or asymptomatic SARS-CoV-2 infection

Rates of any PCR-positive result, irrespective of symptoms, were highest in unvaccinated seronegative HCWs (635 cases), with 85% lower incidence in unvaccinated seropositive HCWs (12 cases, aIRR=0.15 [95%CI 0.08-0.26, p<0.001]). Incidence was reduced by 64% in seronegative HCWs following 1^st^ vaccination (64 cases, aIRR=0.36 [0.26-0.50; p<0.001]) and 90% following 2^nd^ vaccination (2 cases, aIRR=0.10 [0.02-0.38; p<0.001]) (Figures 1C and 2, Tables 2 and S2). Incidence was also 96% lower in vaccinated previously seropositive HCWs (1 case, aIRR=0.04 [0.01-0.27; p=0.001]). As seen above for symptomatic infection, protection from any PCR positive result irrespective of symptoms was lower following first vaccination than if seropositive (p=0.006), but with no evidence of difference between seropositivity and second vaccination (p=0.59).

### PCR-positive results following vaccination

The incidence of PCR-positive results fell from >14 days after the first vaccination for both the Pfizer-BioNTech and Oxford-AstraZeneca vaccines, with similar levels of protection seen up to 42 days post-vaccine (Figure 3). There was an unexpected rise in incidence above baseline levels in the first two weeks following vaccination, which remained to some extent after adjustment (Figure 3B).Considering efficacy against any PCR-positive result >14 days post 1^st^ dose, there was no evidence of a difference by vaccine type following the 1^st^ (heterogeneity p=0.33) or 2^nd^ (p=0.16) dose. Similarly, there was no evidence of difference in PCR-confirmed symptomatic SARS-CoV-2 infection (p=0.21 and p>0.99 respectively).

**Figure 3.**
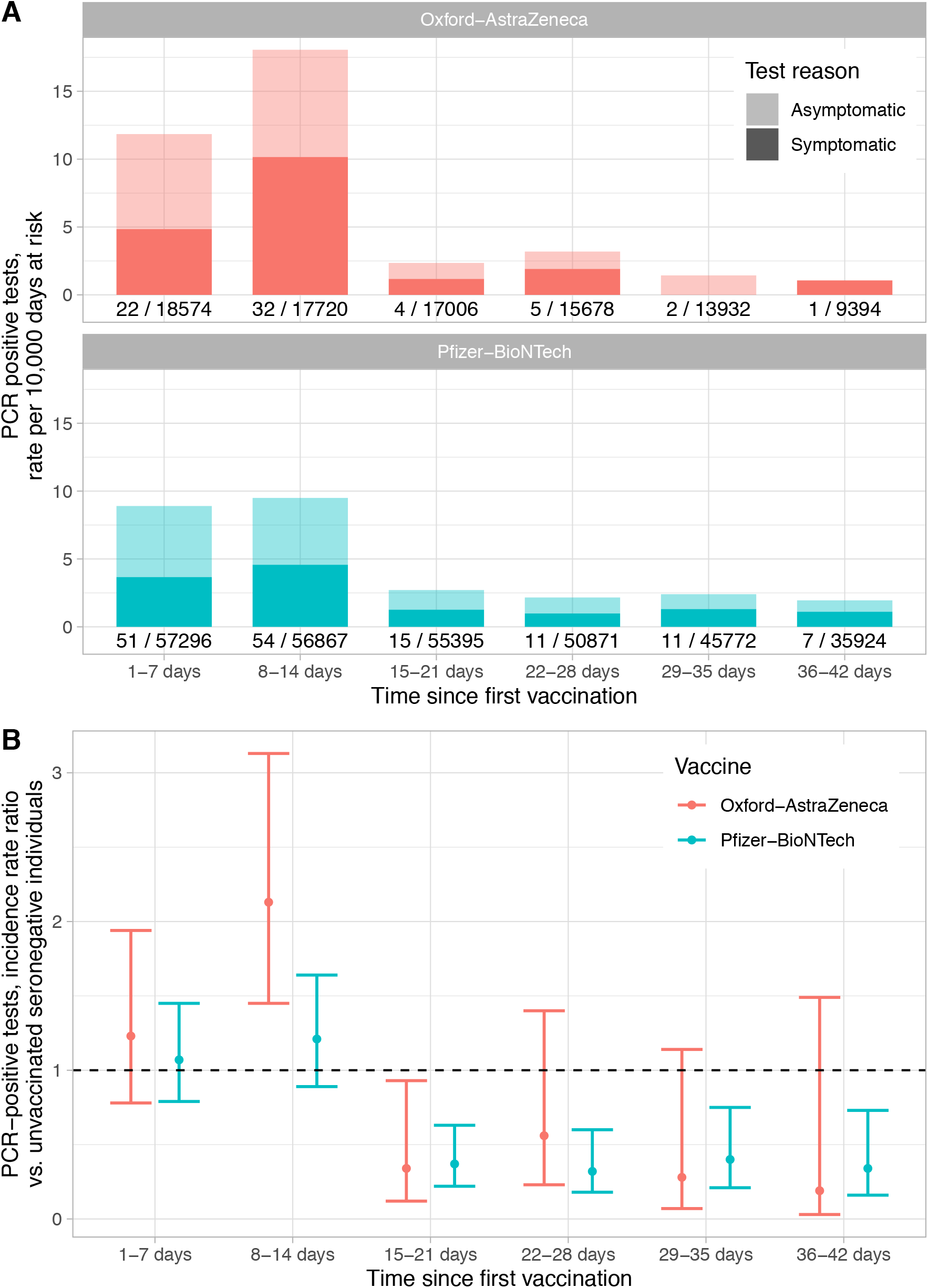
PCR-positive results following first vaccination. Panel A shows observed rates of symptomatic and asymptomatic PCR-positive results; counts and days at risk plotted under each bar. Panel B shows the relative incidence of PCR-positive results by vaccine and days since first vaccine compared to rates in unvaccinated seronegative HCWs. For both plots follow-up is censored if a second vaccination was given.

### Impact of antibody status and vaccination on viral loads

Viral loads were higher, i.e., Ct values lower, in symptomatic infections (median [IQR] Ct 16.3 [13.5-21.7]) compared to asymptomatic screening (20 [14.5-29.5]) (Figure 4A, Kruskal-Wallis p<0.001).Unvaccinated seronegative HCWs had the highest viral loads (Ct: 18.3 [14.0-25.5]), followed by vaccinated previously seronegative HCWs (Ct: 19.7 [15.0-27.5]); unvaccinated seropositive HCWs had the lowest viral loads (Ct: 27.2 [18.8-32.2]) (Figure 4B, overall p=0.06). Combining symptom status and prior-antibody/vaccine status, there was a trend towards pre-vaccine seropositivity and vaccination independently decreasing viral loads, reflected in Ct value increases of 5.7 (95% CI - 0.9,+13.2) and 2.7 (−0.5,+6.8) respectively (Table S4).

**Figure 4.**
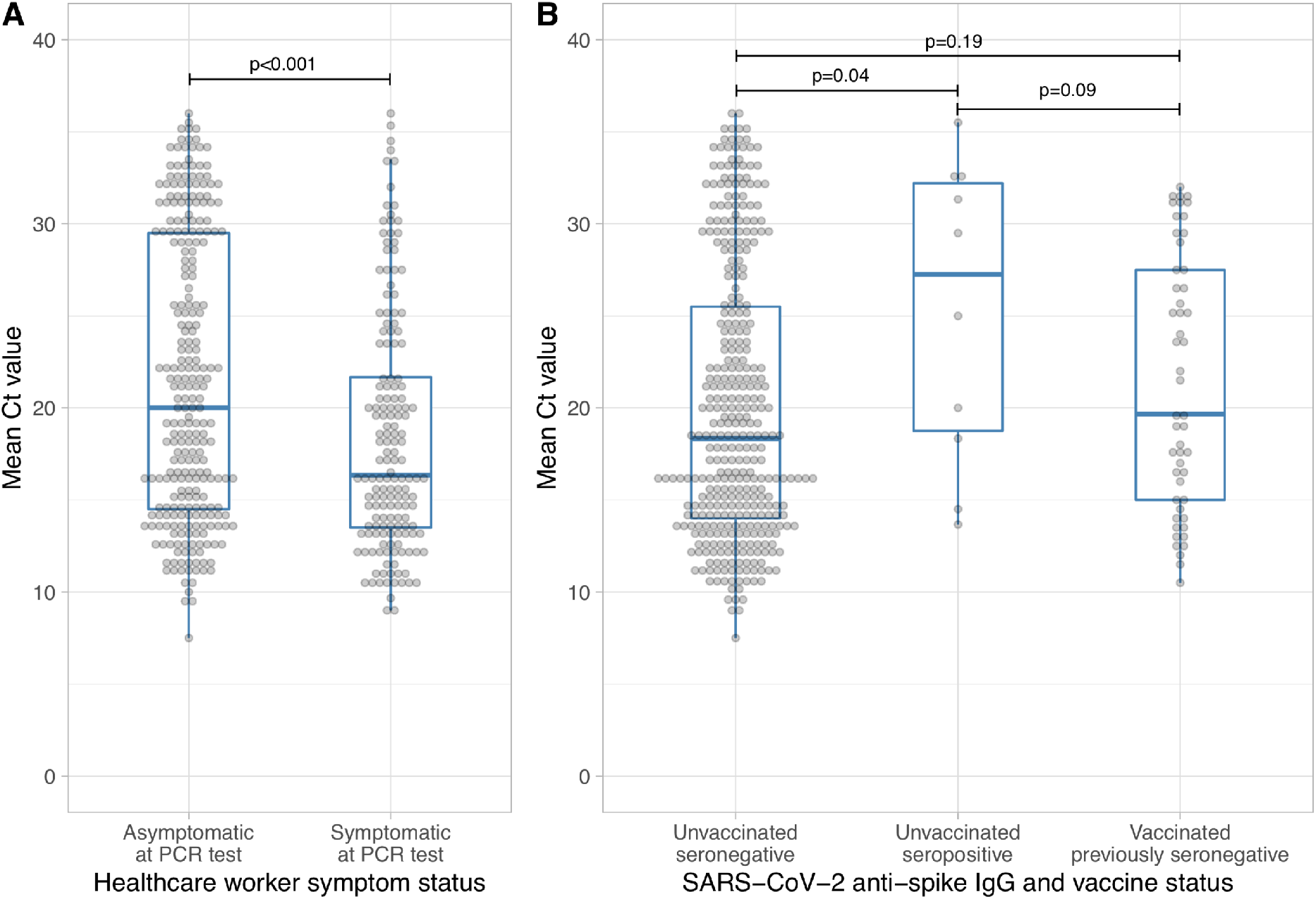
Relationship between SARS-CoV-2 PCR cycle threshold (Ct) values and symptoms (panel A), antibody and vaccine status (panel B). Ct values were available for HCWs tested using the Thermo Fisher TaqPath assay from 16^th^ November 2020 onwards, n=423. The mean per sample Ct value across all detected targets is shown. For panel A, Kruskal-Wallis p<0.001 and for panel B Kruskal-Wallis p=0.06, Wilcoxon rank sum test p values are shown between categories in panel B.

### Incidence of SGTF and B.1.1.7 SARS-CoV-2 infection

From 01-December-2020, SGTF status was determined for 390/463(84%) PCR-positives (with the majority of remaining positive tests undertaken in the community); 258/390(66%) had SGTF. SGTF accounted for 15% of positive PCR results in mid-November-2020, rising to 90% in the second half of January-2021, before declining again (Figure 5A). There was no evidence that SGTF changed the extent of protection against any PCR-positive infection in unvaccinated seropositive HCWs (aIRR vs. non-SGTF, 0.43, [95%CI 0.12-1.52; p=0.19]) or previously seronegative HCWs after a first vaccine (1.13 [0.48-2.63; p=0.78]).

**Figure 5.**
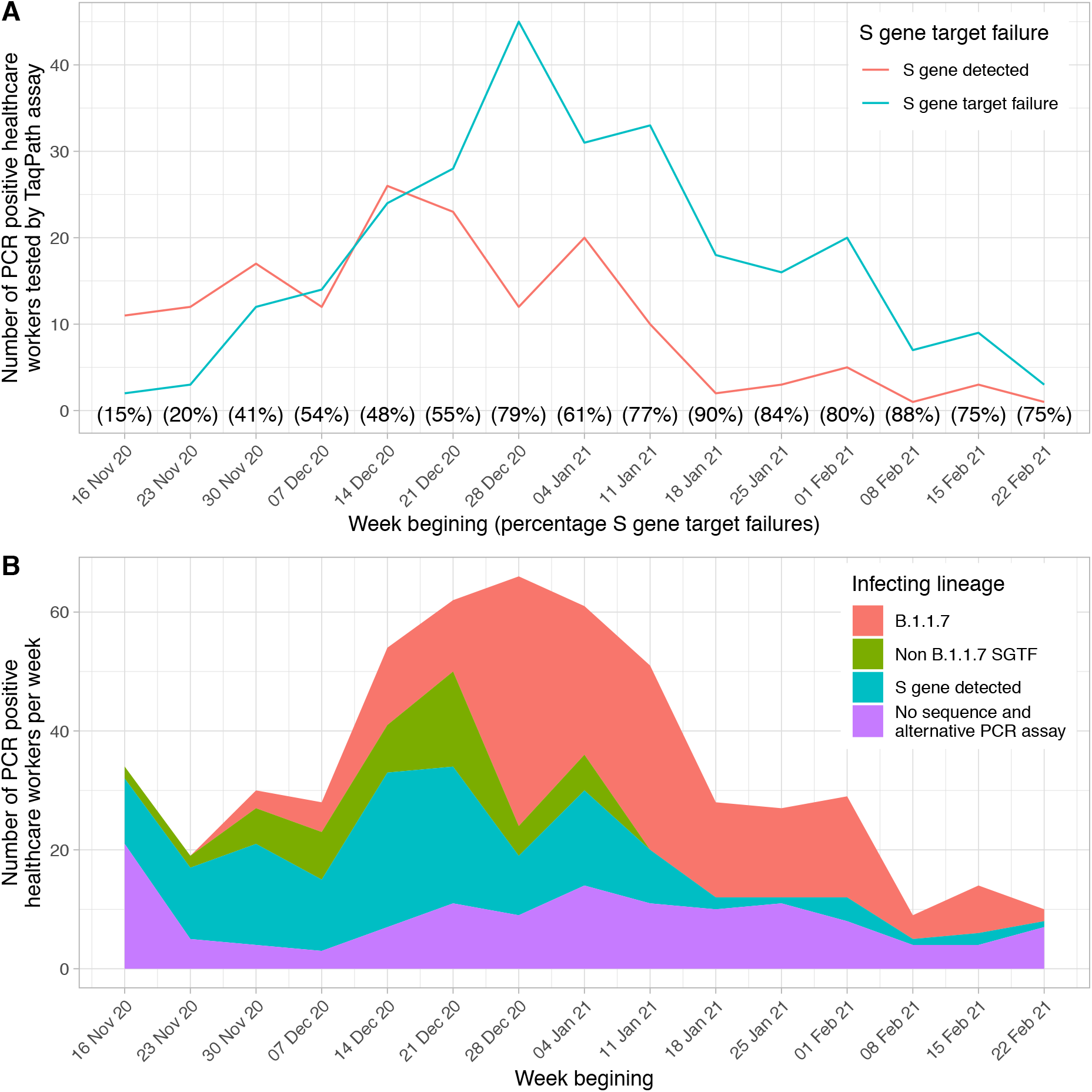
The incidence of S gene target failure (SGTF, panel A) and B.**1**.**1**.**7** (panel B) infection by week of testing. From 16^th^ November 2020 onwards samples from HCWs were routinely processed using the Thermo Fisher TaqPath assay allowing SGTF to be identified, shown in panel A. Sequencing was undertaken of samples processed on other assays as well, hence the larger total in panel B.

We used viral whole-genome sequencing to determine infecting lineages from 01-December-2020 onwards (Table S5): 343/463(74%) were successfully sequenced, 193/343(56%) were B.1.1.7, an additional 19/463(4%) were not sequenced but S-gene positive (i.e. unlikely B.1.1.7) (Figure 5B). There was no evidence that B.1.1.7 changed the extent of protection from any-PCR positive infection in those who were seropositive (aIRR vs non-B.1.1.7=0.40 [95%CI 0.10-1.64; p=0.20]) or following a first vaccine dose (aIRR=1.84 [0.75-4.49; p=0.18). 17% of SGTF was due to a lineage other than B.1.1.7. No other variants of concern (B.1.1.7 with E484K, B.1.351 or P.1) were identified in participating HCWs, in an at-risk period.

## Discussion

In this longitudinal cohort study of HCWs receiving Pfizer-BioNTech and Oxford-AstraZeneca vaccines, vaccination reduced the incidence of PCR-positive symptomatic SARS-CoV-2 infection, with two doses providing similar levels of protection to natural immunity. No symptomatic infections were seen following two vaccine doses and there was a 98% reduction in symptomatic infections in unvaccinated seropositive HCWs. Protection was still afforded >14 days after a single vaccine dose, albeit at lower levels (67% reduction). No vaccinated HCW required hospital admission. Furthermore, vaccination reduced the incidence of any PCR-positivity by 64% and 90% >14 days post-first and second vaccine dose respectively, compared to an 85% reduction post-natural infection. This suggests that both vaccination and previous infection are also likely to reduce transmission. Additionally, there was a trend towards reduced viral loads in re-infected individuals compared to infected seronegative HCWs, with a smaller observed reduction post-vaccination.

The comparable protection offered by seropositivity to two doses of vaccine suggests that the immunoassay used provides an accurate correlate of immunity, which could potentially be used to support individualised relaxation of societal restrictions. Furthermore, where vaccine supplies are limited prioritising seronegative infection-naïve individuals may be appropriate.

Protection following two vaccine doses was comparable to other real-world studies.^6,8^ Protection following a single dose was towards the lower range of previous reports, potentially reflecting occupational exposure in HCWs. Although an unexpected rise in incidence was seen in the first two weeks post vaccination, this time period was excluded from effectiveness calculations. Possible explanations include increased ascertainment of asymptomatic infection due to vaccine-related symptoms leading to testing, behaviour change, acquisition at vaccination facilities, or staff attending for vaccination prompted by high levels of exposure to infected colleagues or patients. A similar rise in incidence was noted in the Israeli mass vaccination programme, attributed to behaviour change post-vaccination.^11,12^

Immunity induced by natural infection and vaccination was robust to lineage, including cases confirmed to be B.1.1.7 by whole-genome sequencing, at least within the power of the study. Sequencing was important to confirm the lineage of SGTF cases: although >99% del69-70 sequences from South-East England were due to B.1.1.7 over this period^20^, locally 17% of SGTF was due to other non-B.1.1.7 lineages. Assuming all SGTF is B.1.1.7 risks misestimating the impact of this lineage on natural and vaccine-induced immunity. This reinforces the need to understand local genomic epidemiology and the reliability of SGTF as a proxy for B.1.1.7 over time. Our results are comparable with the Oxford-AstraZeneca analysis of vaccine efficacy against B.1.1.7 based on a relatively low proportion of successfully sequenced cases (179/499,36%) and no documentation of SGTF status^35^, compared to this study, where PCR and WGS confirmed SGTF/lineage status in 78% cases.

One important finding is that despite universal use of personal protective equipment (gloves, plastic aprons, surgical marks for all patient care and FFP3/N99 masks, gowns and eye protection for aerosol generating procedures), social distancing and use of surgical masks throughout all areas of the hospital, staff working in Covid wards remained at higher risk of SARS-CoV-2 infection independent of vaccine and antibody status. Possible explanations include acquisition from patients with or without subsequent amplification by staff-to-staff spread. Nurses, healthcare assistants and Asian staff were also at higher risk of infection, possibly reflecting both hospital and community-based exposures as we have discussed previously^37^.

One study limitation is that staff working in roles more likely to be exposed to SARS-CoV-2 were initially prioritised for vaccination; these staff were also at the greatest risk of occupationally-acquired SARS-CoV-2 infection. We adjusted for this by including working in a Covid ward and staff roles, but incomplete adjustment could lead to under-estimation of vaccine efficacy. Similarly, vaccinated staff were potentially more likely to be current employees than unvaccinated staff; if unvaccinated seronegative staff left employment this would potentially lead to under-ascertainment of infection in this group. We address this by only including staff using testing and/or vaccination services in the last six months of the study. Testing rates were lower in seropositive HCWs and to a lesser extent following vaccination, leading to under-ascertainment of PCR-positive results in these groups; however, we have previously demonstrated the impact of this is relatively small.^2^ Other limitations include limited power to detect differences in efficacy between vaccines. We were also unable to sequence all PCR-positives, in particular because those with higher Ct values are less likely to generate high-quality sequences, and some samples were not stored, including those processed by community testing facilities. Similar studies will be needed to assess the vaccine effectiveness against other, novel emerging SARS-CoV-2 lineages. Finally, this is a study of HCWs of working age, so findings may not generalise to other settings.

In summary, pooling data from unvaccinated and Pfizer-BioNTech and AstraZeneca vaccinated HCWs, we show that natural infection resulting in detectable anti-spike antibodies and two doses of vaccine both provide robust protection against SARS-CoV-2 infection, including against the B.1.1.7 variant of concern.

## Data Availability

The datasets analysed during the current study are not publicly available as they contain personal data but are available from the Infections in Oxfordshire Research Database (https://oxfordbrc.nihr.ac.uk/research-themes-overview/antimicrobial-resistance-and-modernising-microbiology/infections-in-oxfordshire-research-database-iord/), subject to an application and research proposal meeting the ethical and governance requirements of the Database. Sequence data generated during the study are available from the European Nucleotide Archive under study accession number PRJEB43319.

## Funding

This work was supported by the UK Government’s Department of Health and Social Care. This work was also supported by the National Institute for Health Research Health Protection Research Unit (NIHR HPRU) in Healthcare Associated Infections and Antimicrobial Resistance at Oxford University in partnership with Public Health England (PHE) (NIHR200915), the NIHR Biomedical Research Centre, Oxford, and benefactions from the Huo Family Foundation and Andrew Spokes. The views expressed in this publication are those of the authors and not necessarily those of the NHS, the National Institute for Health Research, the Department of Health or Public Health England. This study is affiliated with Public Health England’s SARS-CoV-2 Immunity & Reinfection EvaluatioN (SIREN) study.

DWE is a Robertson Foundation Fellow and an NIHR Oxford BRC Senior Fellow. SFL is a Wellcome Trust Clinical Research Fellow. DIS is supported by the Medical Research Council (MR/N00065X/1). PCM holds a Wellcome Intermediate Fellowship (110110/Z/15/Z). BDM is supported by the SGC, a registered charity (number 1097737) that receives funds from AbbVie, Bayer Pharma AG, Boehringer Ingelheim, Canada Foundation for Innovation, Eshelman Institute for Innovation, Genome Canada through Ontario Genomics Institute [OGI-055], Innovative Medicines Initiative (EU/EFPIA) [ULTRA-DD grant no. 115766], Janssen, Merck KGaA, Darmstadt, Germany, MSD, Novartis Pharma AG, Pfizer, Sao Paulo Research Foundation-FAPESP, Takeda, and Wellcome. BDM is also supported by the Kennedy Trust for Rheumatology Research. GS is a Wellcome Trust Senior Investigator and acknowledges funding from the Schmidt Foundation. TMW is a Wellcome Trust Clinical Career Development Fellow (214560/Z/18/Z). ASW is an NIHR Senior Investigator.

## Declaration of interests

DWE declares lecture fees from Gilead, outside the submitted work. RJC is a founder shareholder and consultant to MIROBio, outside the submitted work. No other author has a conflict of interest to declare.

## Acknowledgements

We would like to thank all OUH staff who participated in the staff testing program, and the staff and medical students who ran the program. This work uses data provided by healthcare workers and collected by the UK’s National Health Service as part of their care and support. We thank all the people of Oxfordshire who contribute to the Infections in Oxfordshire Research Database. Research Database Team: L Butcher, H Boseley, C Crichton, DW Crook, DW Eyre, O Freeman, J Gearing (community), R Harrington, K Jeffery, M Landray, A Pal, TEA Peto, TP Quan, J Robinson (community), J Sellors, B Shine, AS Walker, D Waller. Patient and Public Panel: G Blower, C Mancey, P McLoughlin, B Nichols.

## References

1. WHO Coronavirus disease (COVID-19) dashboard. COVID 19 Special Issue 10, (2020).

2. Lumley, S. F. et al. Antibody Status and Incidence of SARS-CoV-2 Infection in Health Care Workers. N. Engl. J. Med. (2020) doi:10.1056/NEJMoa2034545.

3. Hanrath, A. T., Payne, B. A. I. & Duncan, C. J. A. Prior SARS-CoV-2 infection is associated with protection against symptomatic reinfection. J. Infect. (2020) doi:10.1016/j.jinf.2020.12.023.

4. Hall, V. et al. Do antibody positive healthcare workers have lower SARS-CoV-2 infection rates than antibody negative healthcare workers? Large multi-centre prospective cohort study (the SIREN study), England: June to November 2020. bioRxiv (2021) doi:10.1101/2021.01.13.21249642.

5. Medicines and Healthcare products Regulatory Agency. MHRA guidance on coronavirus (COVID-19). https://www.gov.uk/government/collections/mhra-guidance-on-coronavirus-covid-19 (2020).

6. Dagan, N. et al. BNT162b2 mRNA Covid-19 Vaccine in a Nationwide Mass Vaccination Setting. N. Engl. J. Med. (2021) doi:10.1056/NEJMoa2101765.

7. Covid-19 vaccine effectiveness surveillance report.

8. Hall, V. J. et al. Effectiveness of BNT162b2 mRNA Vaccine Against Infection and COVID-19 Vaccine Coverage in Healthcare Workers in England, Multicentre Prospective Cohort Study (the SIREN Study). (2021) doi:10.2139/ssrn.3790399.

9. Polack, F. P. et al. Safety and Efficacy of the BNT162b2 mRNA Covid-19 Vaccine. N. Engl. J. Med. 383, 2603–2615 (2020).

10. Report to JCVI on estimated efficacy of a single dose of Pfizer BioNTech (BNT162b2 mRNA) vaccine and of a single dose of ChAdOx1 vaccine (AZD1222).

11. Chodick, G. et al. The effectiveness of the first dose of BNT162b2 vaccine in reducing SARS-CoV-2 infection 13-24 days after immunization: real-world evidence. bioRxiv (2021) doi:10.1101/2021.01.27.21250612.

12. Hunter, P. R. & Brainard, J. Estimating the effectiveness of the Pfizer COVID-19 BNT162b2 vaccine after a single dose. A reanalysis of a study of ‘real-world’ vaccination outcomes from Israel. bioRxiv (2021) doi:10.1101/2021.02.01.21250957.

13. Amit, S., Regev-Yochay, G., Afek, A., Kreiss, Y. & Leshem, E. Early rate reductions of SARS-CoV-2 infection and COVID-19 in BNT162b2 vaccine recipients. Lancet (2021) doi:10.1016/S0140-6736(21)00448-7.

14. Voysey, M. et al. Single-dose administration and the influence of the timing of the booster dose on immunogenicity and efficacy of ChAdOx1 nCoV-19 (AZD1222) vaccine: a pooled analysis of four randomised trials. Lancet (2021) doi:10.1016/S0140-6736(21)00432-3.

15. Voysey, M. et al. Safety and efficacy of the ChAdOx1 nCoV-19 vaccine (AZD1222) against SARS-CoV-2: an interim analysis of four randomised controlled trials in Brazil, South Africa, and the UK. Lancet 397, 99–111 (2021).

16. Effectiveness of first dose of COVID-19 vaccines against hospital admissions in Scotland: national prospective cohort study of 5.4 million people. doi:https://www.ed.ac.uk/files/atoms/files/scotland_firstvaccinedata_preprint.pdf.

17. Walker, A. S. et al. Increased infections, but not viral burden, with a new SARS-CoV-2 variant. bioRxiv (2021) doi:10.1101/2021.01.13.21249721.

18. Davies, N. G. et al. Estimated transmissibility and severity of novel SARS-CoV-2 Variant of Concern 202012/01 in England. bioRxiv (2020) doi:10.1101/2020.12.24.20248822.

19. Volz, E. et al. Transmission of SARS-CoV-2 Lineage B.1.1.7 in England: Insights from linking epidemiological and genetic data. bioRxiv (2021) doi:10.1101/2020.12.30.20249034.

20. Investigation of SARS-CoV-2 variants of concern in England: Technical Briefing 6.

21. Preliminary genomic characterisation of an emergent SARS-CoV-2 lineage in the UK defined by a novel set of spike mutations. https://virological.org/t/preliminary-genomic-characterisation-of-an-emergent-sars-cov-2-lineage-in-the-uk-defined-by-a-novel-set-of-spike-mutations/563 (2020).

22. Graham, M. S. et al. The effect of SARS-CoV-2 variant B.1.1.7 on symptomatology, re-infection and transmissibility. medRxiv 2021.01.28.21250680 (2021).

23. Starr, T. N. et al. Prospective mapping of viral mutations that escape antibodies used to treat COVID-19. Science (2021) doi:10.1126/science.abf9302.

24. Wang, P. et al. Increased Resistance of SARS-CoV-2 Variants B.1.351 and B.1.1.7 to Antibody Neutralization. bioRxiv (2021) doi:10.1101/2021.01.25.428137.

25. Wang, Z. et al. mRNA vaccine-elicited antibodies to SARS-CoV-2 and circulating variants. bioRxiv (2021) doi:10.1101/2021.01.15.426911.

26. Graham, C. et al. Impact of the B.1.1.7 variant on neutralizing monoclonal antibodies recognizing diverse epitopes on SARS-CoV-2 Spike. Cold Spring Harbor Laboratory 2021.02.03.429355 (2021) doi:10.1101/2021.02.03.429355.

27. Shen, X. et al. SARS-CoV-2 variant B.1.1.7 is susceptible to neutralizing antibodies elicited by ancestral Spike vaccines. Cold Spring Harbor Laboratory 2021.01.27.428516 (2021) doi:10.1101/2021.01.27.428516.

28. Skelly, D. T. Vaccine-induced immunity provides more robust heterotypic immunity than natural infection to emerging SARS-CoV-2 variants of concern. doi:10.21203/rs.3.rs-226857/v1.

29. Collier, D. A. et al. Impact of SARS-CoV-2 B.1.1.7 Spike variant on neutralisation potency of sera from individuals vaccinated with Pfizer vaccine BNT162b2. bioRxiv (2021) doi:10.1101/2021.01.19.21249840.

30. Supasa, P. et al. Reduced neutralization of SARS-CoV-2 B.1.1.7 variant by convalescent and vaccine sera. Cell (2021) doi:10.1016/j.cell.2021.02.033.

31. Tada, T. et al. Neutralization of viruses with European, South African, and United States SARS-CoV-2 variant spike proteins by convalescent sera and BNT162b2 mRNA vaccine-elicited antibodies. bioRxiv (2021) doi:10.1101/2021.02.05.430003.

32. Diamond, M. et al. SARS-CoV-2 variants show resistance to neutralization by many monoclonal and serum-derived polyclonal antibodies. Res Sq (2021) doi:10.21203/rs.3.rs-228079/v1.

33. Hu, J. et al. Emerging SARS-CoV-2 variants reduce neutralization sensitivity to convalescent sera and monoclonal antibodies. Cold Spring Harbor Laboratory 2021.01.22.427749 (2021) doi:10.1101/2021.01.22.427749.

34. Xie, X. et al. Neutralization of N501Y mutant SARS-CoV-2 by BNT162b2 vaccine-elicited sera. Cold Spring Harbor Laboratory 2021.01.07.425740 (2021) doi:10.1101/2021.01.07.425740.

35. Emary, K. R. W. et al. Efficacy of ChAdOx1 nCoV-19 (AZD1222) Vaccine Against SARS-CoV-2 VOC 202012/01 (B.1.1.7). (2021) doi:10.2139/ssrn.3779160.

36. Weekes, M. et al. Single-dose BNT162b2 vaccine protects against asymptomatic SARS-CoV-2 infection. Authorea Preprints (2021) doi:10.22541/au.161420511.12987747/v1.

37. Eyre, D. W. et al. Differential occupational risks to healthcare workers from SARS-CoV-2 observed during a prospective observational study. Elife 9, (2020).

38. Lumley, S. F. et al. The duration, dynamics and determinants of SARS-CoV-2 antibody responses in individual healthcare workers. Clin. Infect. Dis. (2021) doi:10.1093/cid/ciab004.

39. National SARS-CoV-2 Serology Assay Evaluation Group. Performance characteristics of five immunoassays for SARS-CoV-2: a head-to-head benchmark comparison. Lancet Infect. Dis. (2020) doi:10.1016/S1473-3099(20)30634-4.

40. Official UK Coronavirus Dashboard. https://coronavirus.data.gov.uk/details/download.

