## Supplementary Material for "An observational cohort study on the incidence of SARS-CoV-2 infection and B.1.1.7 variant infection in healthcare workers by antibody and vaccination status"

#### Group authorship


**Oxford University Hospitals Staff Testing Group:**
**University of Oxford Medical School staff testing team (University of Oxford, Oxford, UK):** Afrah Shibu, Aisling Curtis, Alexandra Mighiu, Ali Manji, Andrey Nezhentsev, Arun Somanathan, Beinn Khulusi, Ben Holloway, Caitlin Rigler, Charis Virgo, Charlotte Fields, Charlotte Lee, Elizabeth Daly, Elizabeth Hatton, Esme Weeks, Euan McGivern, Greta Economides, Hannah Fuchs, Harry Jackson-Smith, Heather Tong, Helen Callard, Helen Clay, Henrietta Davies, Isaac Jarratt Barnham, Ishta Sharma, Jack Wilson, Jocelyn Ward, Joseph Cutteridge, Julia Kotowska, Kirsten Lee, Krupa Ravi, Laura Wilkins, Lottie Cansdale, Lucy Bland, Luiza Farache Trajano, Magdalena Chmura, Maria Lucey, Maria Pikoula, Meirian Evans, Molly Abbott, Morwenna Tamblyn, Oriane Grant, Rebecca Conway-Jones, Ross Toward, Roxanna Abhari, Ruby Wolman, Sara Hosseinzadeh, Sarah Thomas, Tara Madsen, Thomas H Foord, Thomas Johnson, Vimukthi Perera, Zamin Shabir

**University of Oxford staff testing team (University of Oxford, Oxford, UK):** Thomas Christott, George Doherty, Philip W Fowler, Fredrik Karpe, James Kavanagh, Lucas Martins Ferreira, Matt J Neville, Hayleah Pickford, Donal Skelly, Jeremy Swann, Sarah Cameron, Phoebe Tamblin-Hopper, Magda Wolna, Rachael Brown, Denis Volk, Fan Yang-Turner

**Oxford University Hospitals staff testing team (Oxford University Hospitals NHS Foundation Trust, Oxford, UK):** Adrian Bialek, Alison Whitty, Annie Westlake, Barbara Woznika, Bryony Butler, Claudio Ferreira, Danielle Russell, Dawn Pether, Elaine Lawson, Eleanor Ross, Eleni Fragkouli, Elizabeth Sims, Emma Mortimore, Geraldine Shaw, Harriet Mullins, Harriett Caroll, Jane Phillips, Jenny Brown, Jess Ponting, Justyna Szczurkowska, Kim Vilca, Kitty Norris, Louise Holland, Michael Luciw, Michelle Gates, Michelle Layton, Nicola Antonucci, Noemi Bodo, Rebecca Millard, Sara Lyden, Sarah Young, Simran Barot, Vanessa Cox, Victoria Wharton, Zoe Thompson

**Oxford University Hospitals microbiology laboratory (Oxford University Hospitals NHS Foundation Trust, Oxford, UK):** Anne Baby, Jasmine Bastable, Kathryn Cann, Reena Chohan, Josie Clarke, Gabriel Cogorno, Sam Cordy, Georgina Coward, David Crawford-Jones, Sean Crawley, Jack Dobson, Bronte Drummond, Laura Dunn, Caleb Edwin, Simon Evans, Mohamad Fadzillah, Jess Gentry, Sarah Hill, Laura Hobden, Nurul Huda, Gemma Innes, Scott Jarvis, Gerald Jesuthasan, Emma Jones, Anita Justice, Lizzie Kalimeris, Richard Kirton, Nakiah Lashley, Sophie Mason, Alex Mobbs, Ahila Murugathasan, Eleanor Mustoe, Gospel Ngoke, Oliver O'Sullivan, Kimberley Odwin, Jack Oliver, Freyja Pattrick, Claudia Pereira, Simon Perry, Tom Potter, Alexander Prentice, Sophie Ramage, Athena Sanders, Kellyanne Savage, Katherine Shimell, Robin Terry, Emma Thornton, Sue Wareing, Annie Welbourne, Maddison Wheatley

**Oxford University Hospitals Infection, Prevention and Control team (Oxford University Hospitals NHS Foundation Trust, Oxford, UK):** Lisa Butcher, Gabriella D'Amato, Ruth Moroney, Gemma Pill, Lydia Rylance-Knight, Claire Sutton, Claudia Salvagno, Merline Tabirao, Sarah Wright

### Supplementary methods

#### PCR assays

RT-PCR was performed using the Public Health England SARS-CoV-2 assay (targeting the RdRp gene), one of five commercial assays: Abbott RealTime (targeting RdRp and N genes; Abbott, Maidenhead, UK), Altona RealStar (targeting E and S genes; Altona Diagnostics, Liverpool, UK), Cepheid Xpert® Xpress SARS-CoV-2 (targeting N2 and E; Cepheid, California, USA), BioFire® Respiratory 2.1 (RP2.1) panel with SARS-CoV-2 (targeting ORF1ab and ORF8; Biofire diagnostics, Utah, USA), Thermo Fisher TaqPath assay (targeting S and N genes, and ORF1ab; Thermo Fisher, Abingdon, UK) or using the ABI 7500 platform (Thermo Fisher, Abingdon, UK) with the US Centers for Disease Control and Prevention Diagnostic Panel of two probes targeting the N gene.

PCR-positive results from community-based symptomatic testing of Oxford University Hospitals (OUH) healthcare workers (HCWs) forwarded by public health agencies were also included (Thermo Fisher TaqPath assay).

#### Vaccinations

Staff vaccinations provided by the hospital were recorded on an electronic database linked to testing data; staff were also able to supply details of vaccinations from other providers when requesting a symptomatic or asymptomatic PCR test.

#### Whole genome sequencing

Sequencing was attempted in all PCR positive samples that had been stored, regardless of cycle threshold (Ct) value. Samples were sequenced using a multiplex PCR based approach with the ARTIC LoCost protocol and v3 primers[1] using R9.4.1 flow cells (Oxford Nanopore Technologies, Oxford, UK). Consensus sequences were generated using ARTIC fieldbioinformatics v1.2.1.[2] All sequences underwent quality control, requiring >50% consensus genome coverage at ≥20 depth, and agreement between Pangolin[3] and Nextclade v2.3.2[4] assignments of lineage B.1.1.7. Duplicate samples from the same individual were excluded from analysis.

#### Statistical analysis

We used Poisson regression to model incidence of SARS-CoV-2 infection per day-at-risk by study group.

We adjusted for calendar month, self-reported age, sex, ethnicity and staff occupational role and patient contact. We grouped self-reported staff ethnicity into 4 categories, white, Asian, Black, other. Similarly, we grouped occupational roles to ensure groups previously noted to be at higher risk of infection were represented separately, including nurses and healthcare assistants, therapists, porters and domestic staff[5]. To address possible bias introduced by prioritisation of vaccines for those staff most at risk of infection initially, we also adjusted for working on a non-ICU ward caring for Covid patients (previously shown to increase risk[5]), which included those working in acute medical, respiratory and infectious disease specialties. Complete covariate data were available for all HCWs analysed. We fitted separate models for PCR-confirmed symptomatic SARS-CoV-2 infection and any PCR-positive result.

We tested for non-linear effects of age using natural cubic splines with up to 5 knots, choosing the best fitting model (a linear model) based on the Bayesian information criterion. All analyses were performed in R version 4.03, using the splines library for non-linear effects, and the car and multicomp libraries to compare IRRs between follow-up groups.

To investigate differences in protection by vaccine type (i.e. Pfizer-BioNTech, Oxford AstraZeneca), we divided the vaccinated follow-up groups by type and formally tested for heterogeneity by vaccine received. To estimate the onset of protection conferred by vaccination we classified days-at-risk in vaccinated individuals from day 1 post vaccination.

We used stacked Poisson regression[6] to test for variation in the incidence of PCR-positive results with and without SGTF considering only events from 01 December 2020 where S gene PCR results were available. Similarly, for cases with available sequencing data, we identified all B.1.1.7 cases (considering all cases without SGTF to be non-B.1.1.7 cases) and compared incidence of B.1.1.7 vs. non-B.1.1.7 infection by follow-up group using stacked regression. For all stacked models we adjusted for non-SGTF/SGTF and B.1.1.7/non-B.1.1.7 calendar month separately.

For positive samples analysed using the Thermo Fisher TaqPath assay (i.e. the most commonly used assay), we compared cycle threshold (Ct) values between symptomatic and asymptomatic infections and by study follow-up group. We used the mean Ct value per sample across all detected targets. We used multivariable quantile regression (R package quantreg) to analyse the joint impact on Ct value of symptom status, and seropositivity/vaccination.

#### Supplementary tables

| Test reason | Follow-up group | Tests performed | Days at risk | Rate per 10,000 days at risk | Incidence rate ratio vs. unvaccinated seronegative HCWs (95% CI) |
|---|---|---|---|---|---|
| **Asymptomatic** | Unvaccinated seronegative | 42,080 | 2,274,675 | 185 | 1 (reference) |
| **Asymptomatic** | Unvaccinated seropositive | 2,521 | 198,520 | 127 | 0.69 (0.66-0.71) |
| **Asymptomatic** | Vaccinated once, previously seronegative | 4,892 | 289,134 | 169 | 0.91 (0.89-0.94) |
| **Asymptomatic** | Vaccinated twice, previously seronegative | 638 | 39,222 | 163 | 0.88 (0.81-0.95) |
| **Asymptomatic** | Vaccinated, previously seropositive | 563 | 33,709 | 167 | 0.90 (0.83-0.98) |
| **Symptomatic** | Unvaccinated seronegative | 2,470 | 2,274,675 | 10.9 | 1 (reference) |
| **Symptomatic** | Unvaccinated seropositive | 192 | 198,520 | 9.7 | 0.89 (0.77-1.03) |
| **Symptomatic** | Vaccinated, previously seronegative | 242 | 289,134 | 8.4 | 0.77 (0.68-0.88) |
| **Symptomatic** | Vaccinated twice, previously seronegative | 40 | 39,222 | 10.2 | 0.94 (0.69-1.28) |
| **Symptomatic** | Vaccinated, previously seropositive | 30 | 33,709 | 8.9 | 0.82 (0.57-1.17) |

**Table S1. Testing rates by follow-up group**.

| Variable | | Person-days of follow-up | Total HCWs in this follow group | Symptomatic PCR-confirmed infection | | | | | Any PCR-positive result | | | | |
|---|---|---|---|---|---|---|---|---|---|---|---|---|---|
| | | | | Events | Rate per 10,000 person-days | Unadjusted IRR | 95% CI | p value | Events | Rate per 10,000 person-days | Unadjusted IRR | 95% CI | p value |
| **Age** | Age, per 10 year increase | | | | | 0.85 | 0.78 - 0.93 | <0.001 | | | 0.89 | 0.83 - 0.94 | <0.001 |
| **Sex** | Female (Reference) | 2,131,174 | 9,765 | 246 | 1.15 | 1.00 | | | 544 | 2.55 | 1.00 | | |
| | Male | 699,684 | 3,321 | 80 | 1.14 | 0.99 | 0.77 - 1.27 | 0.94 | 169 | 2.42 | 0.95 | 0.80 - 1.12 | 0.53 |
| | Other | 4,402 | 23 | 1 | 2.27 | 1.97 | 0.28 – 14.0 | 0.50 | 1 | 2.27 | 0.89 | 0.13 - 6.30 | 0.91 |
| **Patient facing role** | No (Reference) | 626,307 | 2,888 | 61 | 0.97 | 1.00 | | | 134 | 2.14 | 1.00 | | |
| | Yes | 2,208,953 | 10,221 | 266 | 1.20 | 1.24 | 0.94 - 1.63 | 0.14 | 580 | 2.63 | 1.23 | 1.02 - 1.48 | 0.03 |
| **Covid ward** | Not working in Covid ward | 2,603,522 | 12,019 | 634 | 2.44 | 1.00 | | | 290 | 1.11 | 1.00 | | |
| | Working in non-ICU Covid ward | 231,738 | 1,090 | 80 | 3.45 | 1.43 | 1.02 - 2.02 | 0.04 | 37 | 1.60 | 1.42 | 1.12 - 1.79 | 0.003 |
| **Month** | April - July 2020 (Reference) | 686,122 | 9,691 | 24 | 0.35 | 1.00 | | | 86 | 1.25 | 1.00 | | |
| | August 2020 | 302,581 | 9,892 | 5 | 0.17 | 0.47 | 0.18 - 1.24 | 0.13 | 7 | 0.23 | 0.18 | 0.09 - 0.40 | <0.001 |
| | September 2020 | 301,711 | 10,250 | 5 | 0.17 | 0.47 | 0.18 - 1.24 | 0.13 | 11 | 0.36 | 0.29 | 0.16 - 0.54 | <0.001 |
| | October 2020 | 323,494 | 10,647 | 21 | 0.65 | 1.86 | 1.03 - 3.33 | 0.04 | 38 | 1.17 | 0.94 | 0.64 - 1.37 | 0.74 |
| | November 2020 | 322,268 | 11,028 | 55 | 1.71 | 4.88 | 3.02 - 7.88 | <0.001 | 109 | 3.38 | 2.70 | 2.03 - 3.58 | <0.001 |
| | December 2020 | 319,105 | 11,308 | 94 | 2.95 | 8.42 | 5.38 - 13.2 | <0.001 | 217 | 6.80 | 5.43 | 4.22 - 6.97 | <0.001 |
| | January 2021 | 261,497 | 12,111 | 92 | 3.52 | 10.10 | 6.42 - 15.8 | <0.001 | 184 | 7.04 | 5.61 | 4.34 - 7.26 | <0.001 |
| | February 2021 | 318,482 | 12,695 | 31 | 0.97 | 2.78 | 1.63 - 4.74 | <0.001 | 62 | 1.95 | 1.55 | 1.12 - 2.15 | 0.008 |
| **Follow up group** | Unvaccinated seronegative (Reference) | 2,274,675 | 10,513 | 294 | 1.29 | 1.00 | | | 635 | 2.79 | 1.00 | | |
| | Unvaccinated seropositive | 198,520 | 1,273 | 1 | 0.05 | 0.04 | 0.01 - 0.28 | 0.001 | 12 | 0.60 | 0.22 | 0.12 - 0.38 | <0.001 |
| | Vaccinated once, previously seronegative | 289,134 | 9,711 | 31 | 1.07 | 0.83 | 0.57 - 1.2 | 0.32 | 64 | 2.21 | 0.79 | 0.61 - 1.03 | 0.08 |
| | Vaccinated twice, previously seronegative | 39,222 | 940 | 0 | 0.00 | 0.00 | No events | | 2 | 0.51 | 0.18 | 0.05 - 0.73 | 0.02 |
| | Vaccinated, previously seropositive | 33,709 | 974 | 1 | 0.30 | 0.23 | 0.03 - 1.64 | 0.14 | 1 | 0.30 | 0.11 | 0.01 - 0.76 | 0.03 |
| **Ethnic group** | White (Reference) | 2,062,810 | 9,411 | 197 | 0.96 | 1.00 | | | 455 | 2.21 | 1.00 | | |
| | Asian | 466,176 | 2,157 | 91 | 1.95 | 2.04 | 1.59 - 2.62 | <0.001 | 170 | 3.65 | 1.65 | 1.39 - 1.97 | <0.001 |
| | Black | 106,027 | 532 | 13 | 1.23 | 1.28 | 0.73 - 2.25 | 0.38 | 34 | 3.21 | 1.45 | 1.03 - 2.06 | 0.04 |
| | Other | 200,247 | 1,009 | 26 | 1.30 | 1.36 | 0.90 - 2.05 | 0.14 | 55 | 2.75 | 1.25 | 0.94 - 1.65 | 0.13 |
| **Role** | Other (Reference) | 839,609 | 4,046 | 86 | 1.02 | 1.00 | | | 179 | 2.13 | 1.00 | | |
| | Junior doctor | 165,087 | 942 | 25 | 1.51 | 1.00 | 0.95 - 2.31 | 0.09 | 42 | 2.54 | 1.19 | 0.85 - 1.67 | 0.30 |
| | Senior doctor (Consultant) | 197,533 | 834 | 8 | 0.40 | 0.4 | 0.19 - 0.82 | 0.01 | 21 | 1.06 | 0.5 | 0.32 - 0.78 | 0.003 |
| | Healthcare assistant | 266,636 | 1,263 | 45 | 1.69 | 1.65 | 1.15 - 2.36 | 0.01 | 103 | 3.86 | 1.81 | 1.42 - 2.31 | <0.001 |
| | Nurse | 832,203 | 3,579 | 118 | 1.42 | 1.38 | 1.05 - 1.83 | 0.02 | 248 | 2.98 | 1.4 | 1.15 - 1.69 | <0.001 |

| | | | | | | | | | | | | | |
|---|---|---|---|---|---|---|---|---|---|---|---|---|---|
| | Physio-, occupational or speech/language therapist | 91,033 | 419 | 5 | 0.55 | 0.54 | 0.22 - 1.32 | 0.18 | 20 | 2.20 | 1.03 | 0.65 - 1.64 | 0.90 |
| | Porter, domestic staff | 72,794 | 338 | 7 | 0.96 | 0.94 | 0.43 - 2.03 | 0.87 | 19 | 2.61 | 1.22 | 0.76 - 1.97 | 0.40 |
| | Administrator | 370,365 | 1,688 | 33 | 0.89 | 0.87 | 0.58 - 1.3 | 0.50 | 82 | 2.21 | 1.04 | 0.8 - 1.35 | 0.78 |

**Table S2. Follow-up, events and unadjusted incidence rate ratios (IRRs) by age, sex, month and follow-up group.** The 33 HCWs with undisclosed, trans or other gender are omitted from multivariable regression models as this group as a whole had only 1 PCR-positive result. Note that as follow-up after vaccination occurred during a period of higher incidence overall, univariable IRRs shown are confounded and do not reflect vaccine effectiveness, see Table 1 for adjusted results.

| Variable | | Symptomatic PCR-confirmed infection | | | Any PCR-positive result | | |
|---|---|---|---|---|---|---|---|
| | | Adjusted IRR | 95% CI | p value | Adjusted IRR | 95% CI | p value |
| **Age** | Age, per 10 year increase | 0.92 | 0.84 - 1.02 | 0.11 | 0.99 | 0.99 - 1 | 0.08 |
| **Sex** | Female (reference) | 1.00 | | | 1.00 | | |
| | Male | 1.14 | 0.87 - 1.49 | 0.33 | 1.12 | 0.93 - 1.35 | 0.23 |
| **Patient facing role** | No (reference) | 1.00 | | | 1.00 | | |
| | Yes | 1.04 | 0.75 - 1.46 | 0.80 | 1.10 | 0.88 - 1.39 | 0.40 |
| **Covid ward** | Not working in Covid ward | 1.00 | | | 1.00 | | |
| | Working in non-ICU Covid ward | 1.47 | 1.04 - 2.07 | 0.03 | 1.43 | 1.13 - 1.81 | 0.003 |
| **Month** | April - July 2020 (Reference) | 1.00 | | | 1.00 | | |
| | August 2020 | 0.53 | 0.2 - 1.4 | 0.20 | 0.21 | 0.1 - 0.46 | <0.001 |
| | September 2020 | 0.54 | 0.2 - 1.42 | 0.21 | 0.34 | 0.18 - 0.63 | <0.001 |
| | October 2020 | 2.11 | 1.17 - 3.8 | 0.01 | 1.08 | 0.74 - 1.59 | 0.69 |
| | November 2020 | 5.54 | 3.4 - 9.01 | <0.001 | 3.12 | 2.33 - 4.17 | <0.001 |
| | December 2020 | 9.57 | 6.06 - 15.1 | <0.001 | 6.31 | 4.88 - 8.16 | <0.001 |
| | January 2021 | 15.00 | 9.4 - 23.8 | <0.001 | 8.39 | 6.41 - 11 | <0.001 |
| | February 2021 | 7.05 | 3.87 - 12.9 | <0.001 | 3.81 | 2.6 - 5.59 | <0.001 |
| **Follow up group** | Unvaccinated seronegative (Reference) | 1.00 | | | 1.00 | | |
| | Unvaccinated seropositive | 0.02 | 0 - 0.17 | <0.001 | 0.14 | 0.08 - 0.26 | <0.001 |
| | Vaccinated once, previously seronegative | 0.35 | 0.22 - 0.55 | <0.001 | 0.38 | 0.28 - 0.53 | <0.001 |
| | Vaccinated twice, previously seronegative | No events | | | 0.10 | 0.02 - 0.39 | 0.001 |
| | Vaccinated, previously seropositive | 0.07 | 0.01 - 0.51 | 0.009 | 0.04 | 0.01 - 0.28 | 0.001 |
| **Ethnic group** | White (Reference) | 1.00 | | | 1.00 | | |
| | Asian | 1.99 | 1.54 - 2.56 | <0.001 | 1.65 | 1.37 - 1.98 | <0.001 |
| | Black | 1.15 | 0.65 - 2.02 | 0.64 | 1.33 | 0.94 - 1.88 | 0.11 |
| | Other | 1.34 | 0.89 - 2.02 | 0.16 | 1.27 | 0.95 - 1.68 | 0.10 |
| **Role** | Other (Reference) | 1.00 | | | 1.00 | | |
| | Junior doctor | 1.44 | 0.92 - 2.26 | 0.11 | 1.18 | 0.84 - 1.66 | 0.34 |
| | Senior doctor (Consultant) | 0.54 | 0.26 - 1.14 | 0.11 | 0.63 | 0.39 - 1 | 0.05 |
| | Healthcare assistant | 1.73 | 1.19 - 2.5 | 0.004 | 1.85 | 1.44 - 2.37 | <0.001 |
| | Nurse | 1.49 | 1.1 - 2.02 | 0.01 | 1.48 | 1.2 - 1.82 | <0.001 |
| | Physio-, occupational or speech/language therapist | 0.63 | 0.25 - 1.57 | 0.32 | 1.15 | 0.72 - 1.83 | 0.57 |
| | Porter, domestic staff | 1.22 | 0.55 - 2.68 | 0.62 | 1.60 | 0.98 - 2.6 | 0.06 |
| | Administrator | 0.99 | 0.65 - 1.53 | 0.98 | 1.19 | 0.9 - 1.58 | 0.23 |

**Table S3. Adjusted incidence rate ratios (IRRs) for symptomatic PCR-confirmed SARS-CoV-2 infection and any PCR-positive result (symptomatic or asymptomatic) by antibody and vaccine status including only HCWs participating in asymptomatic screening or symptomatic testing from 01 September 2020 onwards.**

| Variable | Coefficient, i.e. Ct value or change in Ct value | 95% CI lower bound | 95% CI upper bound |
|---|---|---|---|
| **Intercept, i.e., median in unvaccinated seronegative HCWs** | 19.3 | 15.8 | 23.4 |
| **Change in median Ct value if symptomatic** | -3.0 | -6.5 | -0.8 |
| **Change in median Ct value if unvaccinated and seropositive** | +5.7 | -0.9 | +13.2 |
| **Change in median Ct value if vaccinated and previously seronegative** | +2.7 | -0.5 | +6.7 |

**Table S4. Relationship between SARS-CoV-2 PCR cycle threshold (Ct) values and symptoms, antibody and vaccine status.** Multivariable results from median regression. All Ct values were obtained using the Thermo Fisher TaqPath assay.

| | Seronegative unvaccinated n=387 | | Seropositive unvaccinated n=10 | | Vaccinated n=66 | |
|---|---|---|---|---|---|---|
| **n** | Symptomatic 185 | Asymptomatic 202 | Symptomatic 1 | Asymptomatic 9 | Symptomatic 31 | Asymptomatic 35 |
| **Data available** | 129 (70%) | 181 (90%) | 1 (100%) | 8 (89%) | 19 (61%) | 24 (68%) |
| **B.1.1.7** | 65 | 90 | 1 | 2 | 15 | 20 |
| **Not B.1.1.7*** | 64 | 91 | 0 | 6 | 4 | 4 |
| **Failed sequencing**** | 3 (2%) | 20 (10%) | 0 (0%) | 1 (11%) | 0 (0%) | 6 (17%) |
| **Sample not available***** | 53 (29%) | 1 (<1%) | 0 (0%) | 0 (0%) | 12 (39%) | 5 (14%) |

**Table S5. Sequencing and SGTF data availability by analysis subgroup for samples from 01 December 2020 onwards.** Missing data in the symptomatic group tended to be due to community testing samples not being stored and hence not available for sequencing, missing data in the asymptomatic group was more likely to be due to a failed sequencing attempt (related to higher Ct values in this subgroup).

*Includes those sequenced and identified to be another lineage, and those presumed not to be B.1.1.7 as S gene was detected. In the vaccinated group two B.1.1, one B.1.1.240, one B.1.177 and one B.1.177.16 were identified, in the seropositive group, four B.1.177, one B.1.258 and one B.1.36.17.

**Includes those cases with SGTF whose sample was available, but sequence data was not of high enough quality to confirm lineage.

***Includes all community samples not available for sequencing, and some samples tested at Oxford University Hospitals but not stored.

#### References


1. Quick, J. nCoV-2019 sequencing protocol v3 (LoCost). (2020).
2. Loman, N. *et al. Artic-network field bioinformatics: 1.2.1*. (2021). doi:10.5281/zenodo.4441073.
3. *Pangolin software package*.
4. Hadfield, J. *et al.* Nextstrain: real-time tracking of pathogen evolution. *Bioinformatics* **34**, 4121–4123 (2018).
5. Eyre, D. W. *et al.* Differential occupational risks to healthcare workers from SARS-CoV-2 observed during a prospective observational study. *Elife* **9**, (2020).
6. Lunn, M. & McNeil, D. Applying Cox regression to competing risks. *Biometrics* **51**, 524–532 (1995).
